# Associations between Exposure to Perfluoroalkyl Substances with Subsequent Body Composition and Glycemic Responses to Bariatric Surgery

**DOI:** 10.64898/2026.03.30.26349786

**Authors:** Saketh Sankara, Matthew R. Smith, Stephanie M. Eick, Damaskini Valvi, Tasha M. Burley, Douglas I. Walker, Edward Lin, Elizabeth M. Hechenbleikner, Lucia A. Gonzalez Ramirez, Paula-Dene C. Nesbeth, Priyathama Vellanki, Barbara A. Gower, Rob McConnell, Dean P. Jones, Jessica A. Alvarez, Vaia Lida Chatzi, Thomas R. Ziegler

## Abstract

Per- and polyfluoroalkyl substances (PFAS) are chemicals linked to obesity and metabolic dysfunction, but their role in bariatric surgery remains poorly understood. This prospective pilot study examined correlations between plasma PFAS concentrations, body composition, and glycemic measures in adults undergoing bariatric surgery. Thirty-two patients (91% female; 66% Black; mean age 43 years) were enrolled preoperatively; twenty-two completed follow-up at a mean 8.6 months post-surgery. Three PFAS (PFHxS, PFNA, and PFOS) were quantified by plasma liquid chromatography-mass spectrometry; body composition and insulin sensitivity were assessed by dual-energy X-ray absorptiometry and intravenous glucose tolerance testing. At baseline, higher plasma PFNA and PFOS concentrations tracked with lower total lean mass (ρ_s_ = -0.46 and -0.48, respectively) and lean mass index (ρ_s_ = -0.46 and -0.42), and PFNA was inversely correlated with body weight (ρ_s_ = -0.40). No baseline associations were observed with adiposity or glycemic indices. Postoperatively, PFHxS concentrations decreased (median = -1.103 ng/mL, p < 0.001), whereas PFNA and PFOS did not change. Average PFNA was positively correlated with postoperative changes in HOMA-IR (ρ_s_ = 0.51) and total lean mass (ρ_s_ = 0.49). No significant associations were observed for average PFHxS or PFOS. These findings suggest that PFNA and PFOS may be linked to reduced lean tissue at baseline, and that PFNA burden modestly tracks with attenuated metabolic and body composition recovery. In an ANCOVA, baseline PFNA was not significantly associated with postoperative HOMA-IR or total lean mass. Larger, longitudinal studies are needed to clarify how PFAS influence these associations.

## Introduction

Per- and polyfluoroalkyl substances (PFAS) are a ubiquitous class of over 10,000 synthetic compounds found in a wide range of industrial and consumer products^1–3^. PFAS can enter the human body through ingestion of contaminated food and drinking water^4–8^, inhalation of indoor air and dust^4,5,7,8^, and dermal contact with treated products^4,6^. Their highly stable chemical properties, particularly their carbon-fluorine bonds, contribute to their persistence in the environment and accumulation in human tissues^4–10^ where more than 96% of individuals in the US had measurable PFAS concentrations^9^. It is well documented that exposure to PFAS is associated with altered immune^4,10–12^ and thyroid function^4,10,11,13^, liver disease^4,10,11^, lipid and insulin dysregulation^4,10,12,14^, and developmental outcomes^4,10,12^. However, research has increasingly focused on the role of PFAS as obesogens^2,3,6,11,15–21^. These complex findings underscore the importance of understanding PFAS’s impact on human health.

Bariatric surgery for obesity provides an opportunity to investigate the dynamics of PFAS and their potential influence on clinical outcomes^15,22,23^. The intended rapid reduction in body weight and improvement in glycemic control after surgery could be affected by the levels of PFAS in the bloodstream, potentially impacting metabolic pathways and hormonal regulation^2,15,16,23,24^. However, studies that track PFAS levels and physiological outcomes in the bariatric-surgery setting remain sparse^15,24^.

This pilot study addresses this knowledge gap by examining the correlations between plasma PFAS concentrations, insulin resistance/glycemic control, and body composition in a cohort of 32 bariatric surgery patients. We quantified three common PFAS chemicals—perfluorooctanesulfonic acid (PFOS), perfluorohexanesulfonic acid (PFHxS), and perfluorononanoic acid (PFNA)—before and after surgery. Recognizing that these PFAS species function as protein-binding amphiphiles rather than traditional lipophiles^25^, we integrated clinical markers of insulin resistance and body composition to explore correlations with PFAS exposure during the significant physiological shifts and volume contraction induced by bariatric surgery in adults.

We hypothesized that plasma PFOS, PFHxS, and PFNA concentrations may be monotonically associated with clinical outcomes—including body weight, BMI, fat mass and visceral adiposity, lean mass, homeostatic model assessment of insulin resistance (HOMA-IR), acute insulin response to glucose (AIRg), disposition index (DI), and sensitivity index (SI).

## Methods

### Setting and Design

A total of 32 adults scheduled for Roux-en-Y gastric bypass or sleeve gastrectomy were enrolled by feasibility of recruitment between 2018 and 2020 at the Emory Bariatric Center. Eligible participants were adults (age >18 years) with severe obesity (BMI ≥ 40 kg/m², or BMI ≥ 35 kg/m² with at least one obesity-related comorbidity: hypertension, type 2 diabetes mellitus, dyslipidemia, or obstructive sleep apnea). Exclusion criteria included prior gastric bypass or gastrectomy, history of type 1 diabetes mellitus, active or chronic infectious or inflammatory disease, active cancer, alcoholism, or severe psychosocial disorder within the prior year. Each participant attended a baseline visit in the Georgia Clinical Translational Science Alliance (Georgia CTSA) Clinical Research Center (CRC) at Emory University Hospital, in Atlanta GA within two months prior to surgery. Twenty-two of these individuals underwent bariatric surgery and returned for a follow-up visit to the CRC an average of 8.6 months later (Standard error: 0.6 months). Both CRC visits included plasma sample collection for the measurement of PFAS, dual energy X-ray absorptiometry (DXA) scans for body-composition and a frequently sampled intravenous glucose tolerance test (IVGTT)^26^.

Plasma aliquots were analyzed by liquid chromatography and mass spectrometry (LC-MS) for PFOS, PFHxS, and PFNA in the Emory Clinical Biomarkers Laboratory. These PFAS species were confirmed by comparing their LC-MS retention times (± 30 seconds) and mass-to-charge (m/z) ratios (± 5 ppm) to those of their respective standards in an in-house library of metabolites previously validated using ion-dissociation tandem mass spectrometry (MS/MS). Within-assay analytical precision, assessed from triplicate injections of QC/reference samples and summarized as the median across QC groups, yielded CVs of 6.4% for PFOS, 45.0% for PFHxS, and 70.0% for PFNA (n=8 QC groups). Metabolite score-based annotation was conducted using an adapted scoring criteria^27^. The limits of detection (LOD) ranged from 0.03 to 0.1 ng/mL, % < LOD provided in the supplement (Table S1). Serum PFAS concentrations below LOD were imputed with LOD / √2.28

This study was conducted according to the guidelines laid down in the Declaration of Helsinki, and all procedures involving human participants were approved by the Institutional Review Board at Emory University School of Medicine (IRB Protocol No. 00102564). All participants gave written informed consent.

### Assessment of Body Composition

We calculated body mass index (BMI, kg/m²) using measured weight and height obtained during the pre-surgery and follow-up post-surgery visits in the CRC. All body composition outcomes, including regional percent fat, total fat mass (kg), visceral fat mass (kg), fat mass index, total lean mass (kg), and lean mass index, were assessed at baseline and follow-up visits by DXA (GE Lunar iDXA, GE Healthcare) at the CRC. Fat mass index and lean mass index were calculated by dividing the respective total mass values (kg) by height squared (m²).

### Assessment of Metabolic Markers

Plasma was obtained and stored at –80 °C until analysis. Glucose and insulin concentrations were determined at the UAB Diabetes Research Center. The MINMOD computer program (Millennium version, © Richard N. Bergman^29^) was used to obtain the SI. The software computed the AIRg as the incremental insulin area under the curve from 0–10 min after glucose injection, using the trapezoidal rule. DI, a composite indicator of insulin secretion and sensitivity, was determined as the product of SI and AIRg^29,30^.

### Measurement of PFAS in Plasma and Reference Standardization

The plasma samples were prepared for LC-MS analysis as previously outlined^31,32^. Full details of PFAS determinations are in the supplemental material. PFHxS, PFNA, and PFOS previously confirmed by MS/MS were converted to plasma concentrations using a reference standardization for quantification protocol.^33^

### Statistical Analyses

Descriptive statistics for all study variables were compiled in SAS Studio (2024, SAS Institute Inc.). The three PFAS levels among participants were markedly skewed; therefore, we log transformed the PFAS levels and used non-parametric correlations with the outcome variables.

Boxplots were generated in GraphPad Prism 9.50 to visualize pre- and post-surgical distributions of each PFAS. To formally test whether concentrations differed before and after surgery within individuals, Wilcoxon signed-rank tests were applied. For subsequent analyses, we used unadjusted Spearman rank correlations with 95% confidence intervals to quantify monotonic relationships between PFAS concentrations and relevant outcomes. Assuming a two-sided α = 0.05 and 80% power, with n = 32 and 3 PFAS exposures (PFOS, PFNA, PFHxS) evaluated across 12 outcomes (weight, BMI, regional % fat, total fat mass, visceral fat mass, fat mass index, total lean mass, lean mass index, HOMA-IR, AIRg, DI, SI), our study can detect correlations as low as |ρ|=0.48. Correlations were estimated separately for baseline PFAS with baseline outcomes and average PFAS burden (the geometric mean of the baseline and postoperative PFAS values) with corresponding changes in body composition and glycemic indices. For our main longitudinal models, participants entirely missing postoperative plasma PFAS were excluded utilizing complete-case analysis for the exposure variable. For missing secondary clinical outcomes, we utilized pairwise deletion to maximize the use of available data. P-values less than 0.05 were considered statistically significant. To mitigate potential confounding related to initial clinical severity, we conducted an exploratory analysis using multivariable linear regression (ANCOVA). Each postoperative outcome was modeled as the dependent variable, with baseline log-transformed PFAS and the corresponding baseline clinical value entered as predictors. No additional covariates were included because of the small sample size and exploratory study design. These analyses were performed in R 4.5.2 (R Foundation for Statistical Computing), and results are presented as forest plots in the Supplementary Material.

## Results

### Cohort Characteristics

We analyzed 32 adults (91% women; mean ± SD age = 42.9 ± 14.0 y) at the time of bariatric surgery and re-evaluated 22 of these participants a median of eight months later (Table 1). Specifically, 6 participants were lost to study follow-up due to the COVID-19 pandemic, and 4 participants completed a follow-up clinic visit but lacked complete postoperative plasma PFAS data. Among the final 22 participants analyzed postoperatively, 2 were missing postoperative IVGTT data but were retained for all other body composition and metabolic analyses. At baseline, most participants self-identified as Black or African-American (66%). Among the follow-up cohort, 68% received sleeve gastrectomy and 32% received Roux-en-Y gastric bypass (Table 1).

**Table 1.**
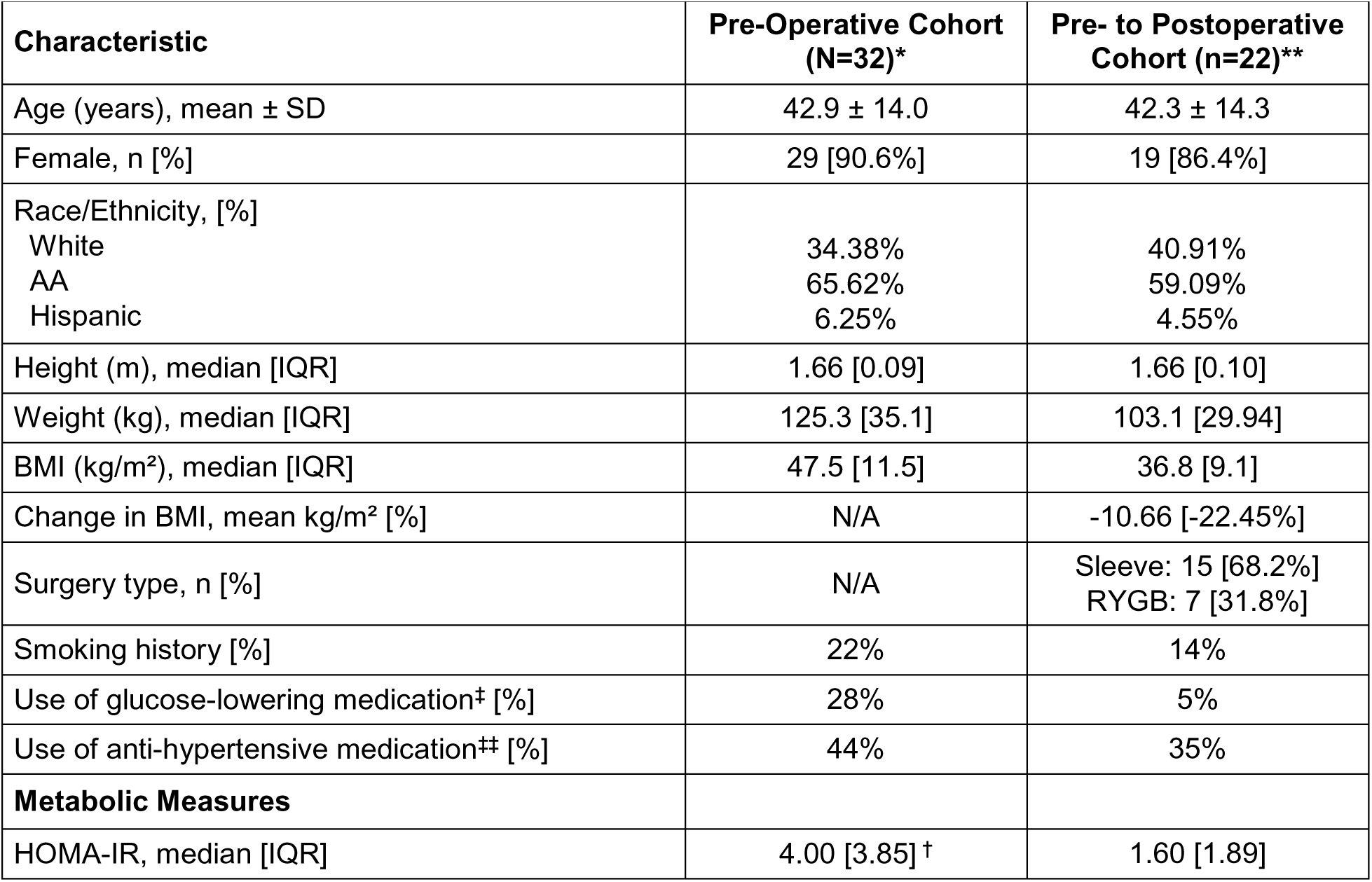
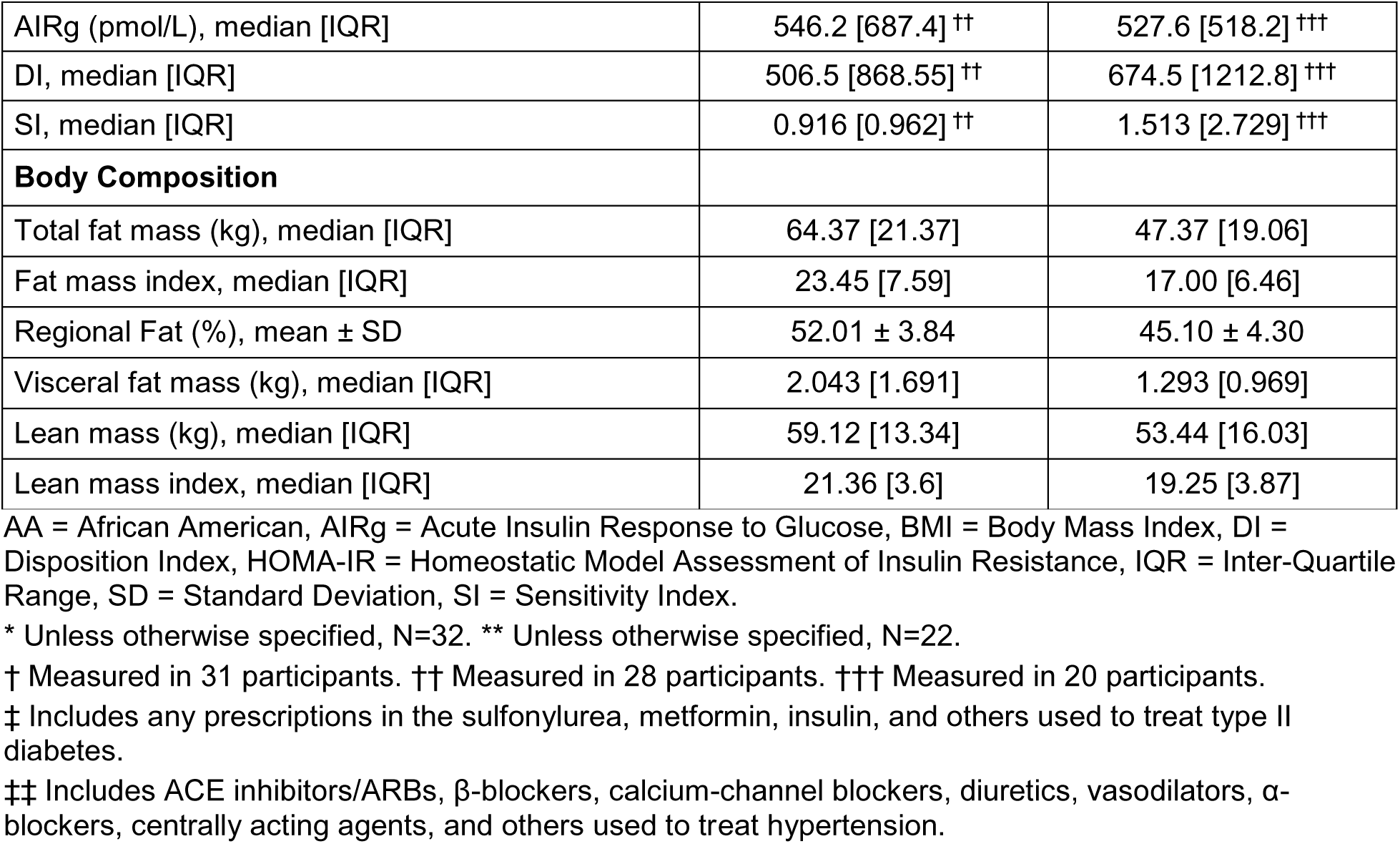
Baseline and Pre- to Postoperative Demographic Characteristics.

At baseline the cohort exhibited severe obesity (median BMI = 47.5 kg m⁻ ²). By follow-up, median BMI had fallen to 36.8 kg m⁻ ² kg—an average BMI reduction of 10.7 kg m⁻ ² (-22.45%). DXA-derived body composition showed similar changes after bariatric surgery: Total fat mass declined from 64.4 kg to 47.4 kg (-26%), visceral fat mass fell by 0.75 kg (-36.7%), and lean mass decreased modestly (59.1 kg to 53.4 kg), representing a 9.9% fall in lean mass index (Table 1).

Baseline insulin resistance was pronounced (median HOMA-IR = 4.00). Postoperatively, HOMA-IR improved to 1.60, while insulin sensitivity (SI) rose 65.2%. β-cell function as measured by DI improved 33.2% post-surgery (Table 1).

### Baseline PFAS Concentrations and Correlations with Baseline Body Composition and Glycemic Indexes

The analysis (Table 2) revealed significant inverse correlation between PFNA levels and preoperative body weight (ρs = -0.40). There were no significant correlations between PFHxS, PFNA, and PFOS with BMI, regional percent fat, total fat mass, fat mass index, or visceral fat mass. However, there were significant inverse correlations with both PFNA and PFOS and total lean mass (ρs = -0.46 and -0.48, respectively). There were also significant inverse correlations with both PFNA and PFOS and lean mass index (ρs = -0.46 and -0.42, respectively) (Table 2).

**Table 2.**
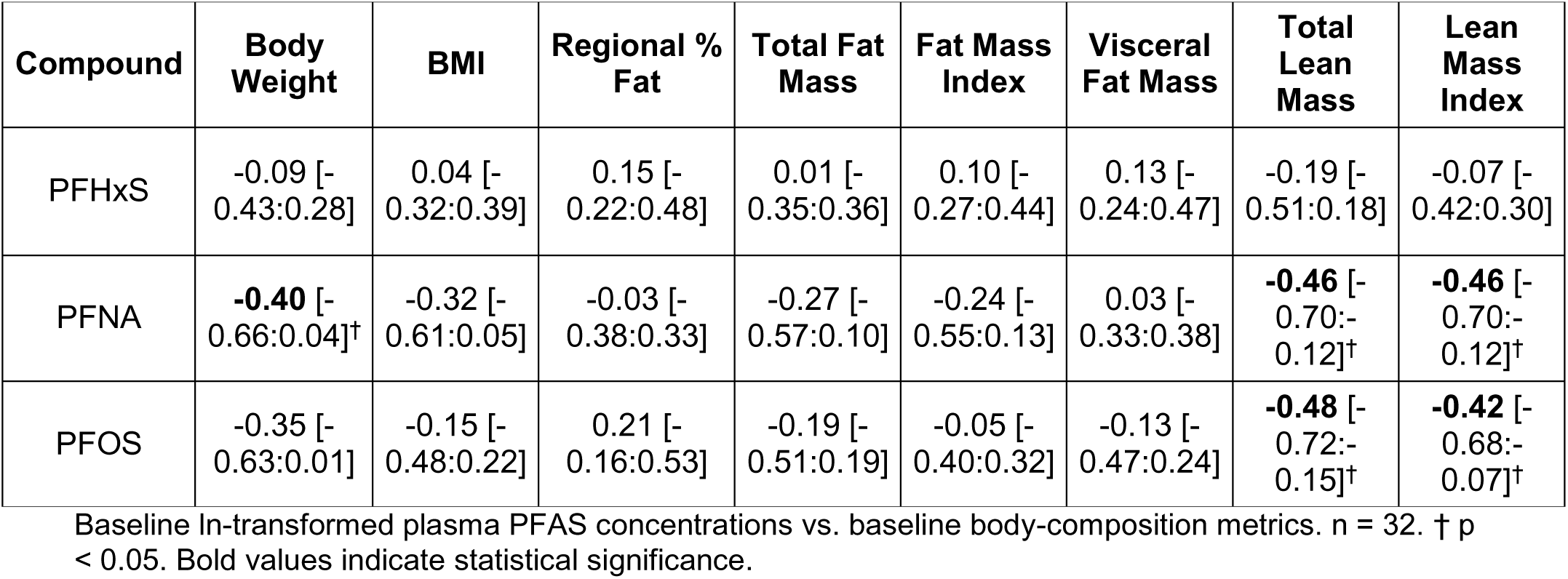
Correlation of Baseline Plasma PFAS Concentrations with Baseline Body Composition (ρ, [95% CI])

| Compound | Body Weight | BMI | Regional % Fat | Total Fat Mass | Fat Mass Index | Visceral Fat Mass | Total Lean Mass | Lean Mass Index |
| --- | --- | --- | --- | --- | --- | --- | --- | --- |
| PFHxS | -0.09 [-0.43:0.28] | 0.04 [-0.32:0.39] | 0.15 [-0.22:0.48] | 0.01 [-0.35:0.36] | 0.10 [-0.27:0.44] | 0.13 [-0.24:0.47] | -0.19 [-0.51:0.18] | -0.07 [-0.42:0.30] |
| PFNA | **-0.40** [-0.66:0.04]<sup>†</sup> | -0.32 [-0.61:0.05] | -0.03 [-0.38:0.33] | -0.27 [-0.57:0.10] | -0.24 [-0.55:0.13] | 0.03 [-0.33:0.38] | **-0.46** [-0.70:-0.12]<sup>†</sup> | **-0.46** [-0.70:-0.12]<sup>†</sup> |
| PFOS | -0.35 [-0.63:0.01] | -0.15 [-0.48:0.22] | 0.21 [-0.16:0.53] | -0.19 [-0.51:0.19] | -0.05 [-0.40:0.32] | -0.13 [-0.47:0.24] | **-0.48** [-0.72:-0.15]<sup>†</sup> | **-0.42** [-0.68:-0.07]<sup>†</sup> |

Baseline ln-transformed plasma PFAS concentrations vs. baseline body-composition metrics. n = 32. <sup>†</sup> p < 0.05. Bold values indicate statistical significance.

As shown in Table 3, there were no significant correlations between PFHxS, PFNA, and PFOS with HOMA-IR, AIRg, DI, or SI.

**Table 3.** Correlation of Baseline Plasma PFAS Concentrations with Baseline Glycemic Indices (ρ, [95% CI])

| Compound | HOMA-IR | AIRg | DI | SI |
| --- | --- | --- | --- | --- |
| PFHxS | -0.12 [-0.46:0.25] | 0.11 [-0.27:0.46] | 0.15 [-0.23:0.49] | -0.18 [-0.52:0.20] |
| PFNA | 0.02 [-0.34:0.37] | 0.22 [-0.16:0.54] | 0.12 [-0.25:0.47] | -0.10 [-0.45:0.27] |
| PFOS | 0.03 [-0.33:0.38] | -0.06 [-0.42:0.31] | -0.09 [-0.44:0.28] | -0.03 [-0.38:0.33] |

Baseline ln-transformed plasma PFAS concentrations vs. glycemic indices. HOMA-IR: n=31; AIRg, DI, SI: n=28. No correlations reached statistical significance at p < 0.05.

### Plasma PFAS Characteristics

The results (Figure 1) demonstrate a heterogeneous, species-specific response to these bariatric surgical interventions. We observed a significant decrease in PFHxS levels across the cohort (p<0.001), with a median within-subject change (calculated as the median of individual differences between pre- and postoperative values) of -1.103 ng/mL (Figure 1A). Conversely, changes in PFNA were not statistically significant (p=0.18) (Figure 1B). For PFOS, which exhibited the highest baseline concentrations, the observed median decrease also failed to reach statistical significance (p=0.19) (Figure 1C). Summary statistics of these findings are provided in Supplemental Information Table S1.

**Figure 1.**
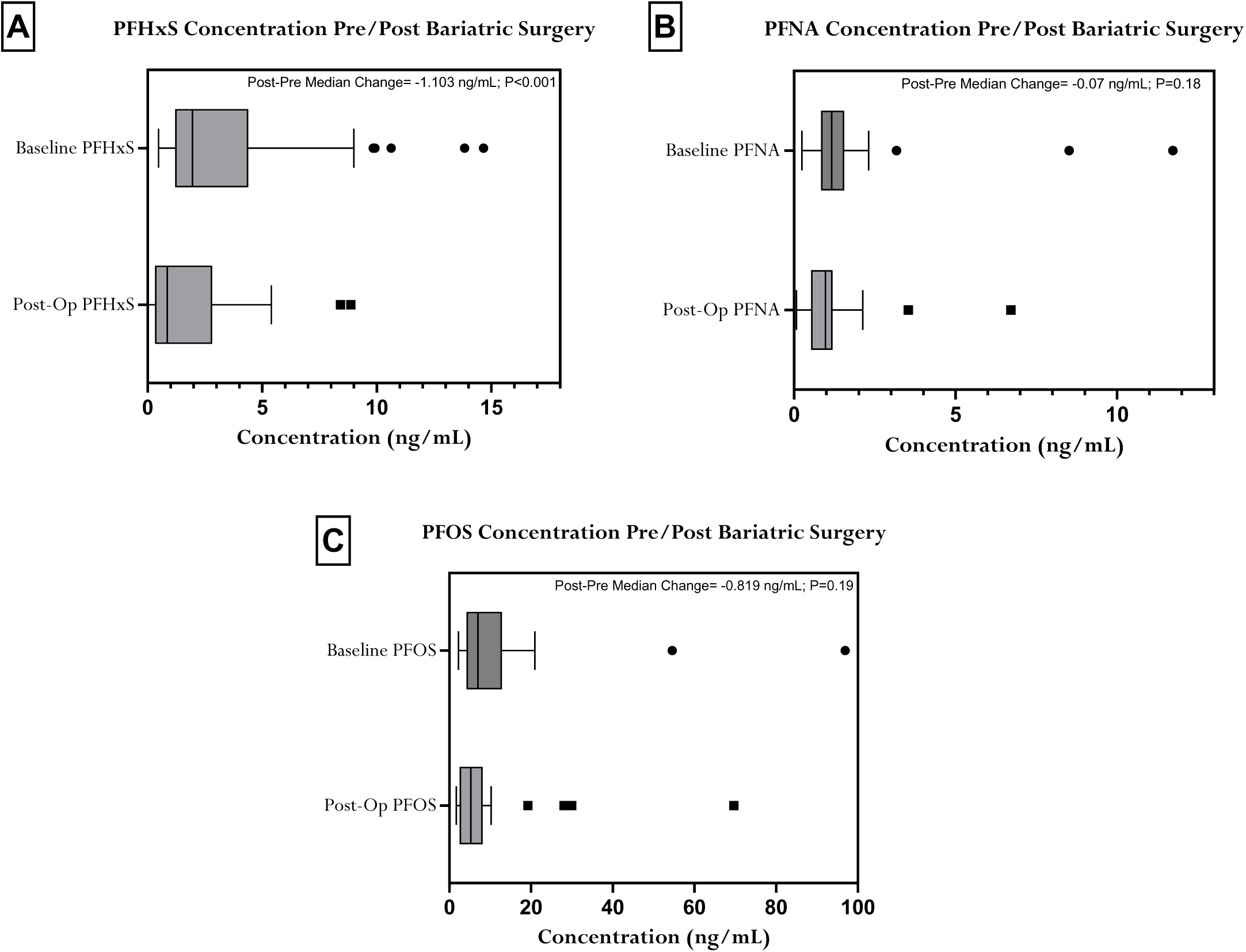
Decline of Plasma PFAS Concentrations after Bariatric Surgery. Box plots compare pre-and postoperative concentrations for (A) PFHxS (n=22), (B) PFNA (n=22), and (C) PFOS (n=22), respectively. Overall, plasma concentrations of PFHxS decreased significantly in the post-surgery follow up (p<0.001), while the decreases observed for PFNA and PFOS were not statistically significant.

### Average PFAS Concentrations and Correlations with Change in Body Composition and Glycemic Indexes Following Bariatric Surgery

To examine the relationship between average PFAS burden and postoperative changes in body composition and glycemic measures, we analyzed data from the 22 participants with complete follow-up data (Tables 4 & 5). There were no statistically significant associations between average PFHxS or PFOS concentrations and measures of adiposity, lean body mass, or glycemic control. However, average PFNA concentrations showed a significant positive correlation with the change in HOMA-IR (ρs = 0.51) (Table 5). Additionally, average PFNA was significantly positively correlated with the change in total lean mass (ρs = 0.49) (Table 4). To further assess these findings, we performed supplementary ANCOVA models in which each postoperative outcome was regressed on baseline log-transformed PFAS and the corresponding baseline clinical value (Figure S1). In these exploratory models, baseline PFNA was not significantly associated with postoperative HOMA-IR or postoperative total lean mass after accounting for the corresponding baseline values.

**Table 4.** Correlation of Average Plasma PFAS Concentrations with the Change in Body Composition after Bariatric Surgery (ρ, [95% CI])

| Compound | Body Weight | BMI | Regional % Fat | Total Fat Mass | Fat Mass Index | Visceral Fat Mass | Total Lean Mass | Lean Mass Index |
| --- | --- | --- | --- | --- | --- | --- | --- | --- |
| PFHxS | -0.16 [-0.55:0.30] | -0.23 [-0.60:0.22] | -0.14 [-0.54:0.31] | -0.15 [-0.55:0.31] | -0.22 [-0.60:0.22] | -0.04 [-0.46:0.40] | -0.17 [-0.56:0.28] | -0.13 [-0.54:0.33] |
| PFNA | 0.22 [-0.23:0.60] | 0.13 [-0.32:0.53] | -0.12 [-0.53:0.33] | 0.12 [-0.33:0.53] | 0.06 [-0.39:0.48] | 0.17 [-0.29:0.56] | **0.49** [0.07:0.76]<sup>†</sup> | 0.36 [-0.08:0.68] |
| PFOS | 0.16 [-0.30:0.55] | 0.08 [-0.36:0.49] | -0.22 [-0.60:0.23] | 0.08 [-0.37:0.49] | 0.01 [-0.42:0.44] | -0.06 [-0.47:0.39] | 0.22 [-0.23:0.60] | 0.10 [-0.35:0.51] |

Average (geometric mean) ln-transformed plasma PFAS concentrations vs. change in body-composition metrics. n = 22. <sup>†</sup> p < 0.05. Bold values indicate statistical significance.

**Table 5.** Correlation of Average Plasma PFAS Concentrations with the Change in Glycemic Indices (ρ, [95% CI])

| Compound | HOMA-IR | AIRg | DI | SI |
| --- | --- | --- | --- | --- |
| PFHxS | 0.17 [-0.28:0.56] | 0.02 [-0.43:0.46] | -0.09 [-0.51:0.38] | -0.23 [-0.61:0.22] |
| PFNA | **0.51** [0.10:0.77]<sup>†</sup> | -0.01 [-0.45:0.44] | -0.18 [-0.58:0.29] | -0.34 [-0.67:0.10] |
| PFOS | 0.27 [-0.18:0.63] | -0.20 [-0.59:0.27] | -0.29 [-0.64:0.16] | -0.24 [-0.62:0.22] |

Average (geometric mean) ln-transformed plasma PFAS concentrations vs. change in glycemic indices. HOMA-IR: n=22; AIRg, DI, SI: n=20. <sup>†</sup> p < 0.05. Bold values indicate statistical significance.

## Discussion

Our goal in this pilot study was to better understand the role of PFAS as obesogens in a bariatric surgery setting. At baseline, higher PFNA and PFOS concentrations tracked with lower lean body mass and lean mass index. While PFNA also showed an inverse relationship with body weight, we observed no clear associations between the other PFAS chemicals and adiposity or glycemic indices. Overall, bariatric surgery was accompanied by a species-specific shift in plasma PFAS: PFHxS levels declined, whereas PFNA and PFOS remained largely unchanged. Using the average (geometric mean) of pre- and postoperative concentrations as an index of overall PFAS exposure, we found that higher PFAS burden did not clearly correspond to most changes in body composition or glycemic indices. Average PFNA was positively correlated with changes in HOMA-IR and total lean mass, whereas no significant associations were observed for average PFHxS or PFOS. Taken together, these findings suggest that while pre-surgical PFAS exposures—particularly PFNA and PFOS—may be linked to reduced lean tissue, PFNA burden may also modestly track with attenuated metabolic and compositional recovery after surgery. However, in supplementary ANCOVA models using baseline PFAS and the corresponding baseline clinical values, baseline PFNA was not significantly associated with postoperative HOMA-IR or total lean mass.

In our pilot study, PFHxS was the only PFAS that declined following bariatric surgery. These findings extend the sparse bariatric surgery literature that has largely emphasized pollutant mobilization and PFAS trajectories in adolescents^15^. In that study, from the Teen-LABS consortium, the authors observed a small, but statistically significant, decrease in PFHxS, PFOS, and PFNA concentrations over 36 months from bariatric surgery (∼70% RYGB)^15^. In contrast, the absence of significant declines for PFNA and PFOS in our cohort likely reflects limited statistical power within our smaller follow-up sample and shorter observation window. However, reports on PFHxS trajectories after surgery have been inconsistent. For example, a Norwegian cohort study observed significant decreases across multiple plasma PFAS, including PFHxS, one year postoperatively^34^, whereas a Finnish study noted a modest increase in serum PFHxS over a similar interval despite minor reductions in other compounds^35^. In contrast, some cohorts have found serum PFAS concentrations to remain stable across the first postoperative year, even as other lipophilic persistent organic pollutants increased markedly, underscoring how chemical properties and cohort context can shape the patterns observed^7^. These divergent findings may reflect compound-specific toxicokinetics, such as the longer half-life and stronger protein binding of PFHxS^9^, or its lack of correlation with circulating lipid and lipoprotein profiles^36^, as well as differences in demographics, and surgical follow-up windows across studies. Taken together, our observation that PFHxS behaved differently from other PFAS suggests that future work should clarify whether PFHxS responds uniquely to weight-loss surgery or whether the apparent variability is a function of study design and sample characteristics.

In our cohort, higher baseline PFNA and PFOS concentrations were linked to indices of lower lean mass. Prior findings in other populations support a similar pattern; in longitudinal data from the Project Viva cohort^37^, children (not undergoing bariatric surgery) with higher plasma PFOS had less accrual of lean mass from mid-childhood to early adolescence. Additionally in that study, higher plasma PFNA was associated with greater accrual of visceral fat mass^37^. Conversely, analyses from a Swedish study of overweight middle-aged adults (not undergoing bariatric surgery) did not identify consistent associations between serum PFOS or PFNA and lean mass, although PFNA did show inverse relationships with several measures of adiposity among women rather than with lean compartments^2^. Also, analyses of NHANES 2011–2018 found that serum concentrations of both PFNA and PFOS were inversely associated with lean mass index in U.S. adults, with associations most evident among women and individuals with obesity, while patterns were weaker or even positive in adolescents^20^. Of note, sex and race/ethnicity have been reported to modify PFAS exposure and body composition outcomes^9,20^. Larger, longitudinal investigations are needed to determine whether and how PFAS may contribute to reduced lean mass, or if observed correlations are markers of other physiological processes.

Baseline PFNA concentrations were inversely associated with body weight in our cohort. Analyses from a large study of midlife women over nearly two decades, did not identify significant associations between serum PFNA and trajectories of body weight or adiposity^38^. Also, a recent cross-sectional study showed that PFNA was inversely associated with several markers of adiposity, including BMI, waist circumference, and fat depots, with these patterns most evident among women^2^. Therefore, there appears to be heterogeneity in the associations between PFAS species and body weight/composition depending on the metric used, population characteristics, and the analytic approach.

When evaluating average plasma PFAS concentrations in relation to postoperative changes, PFNA exhibited significant positive correlations with both HOMA-IR and total lean mass. Participants with higher average PFNA concentrations experienced a smaller postoperative reduction in HOMA-IR, interpreted as reduced improvement in insulin resistance. Although our small sample size precludes any causal conclusion, the direction of this association is consistent with prior epidemiologic studies in non-bariatric surgery patients that have linked PFNA exposures to elevated HOMA-IR, fasting insulin, and impaired glucose regulation in adults^16,17^. Additionally, we observed that higher average PFNA was significantly correlated with a smaller loss of total lean mass. However, rather than indicating a protective effect against muscle loss, this observation may reflect an effect of our baseline findings: individuals with higher PFNA exposure already presented with significantly lower lean mass prior to surgery, which means less lean tissue to lose after bariatric surgery. Impairment of lean mass accrual is consistent with previous literature evaluating PFAS during weight loss^37,39^. Ultimately, while these preliminary data suggest that specific PFAS burden may influence the extent of metabolic and compositional recovery, larger, adequately powered longitudinal studies are needed to test these relationships and rule out residual confounding.

## Strengths and Limitations

This study has several strengths. It is, to our knowledge, one of the earliest investigations of plasma PFAS dynamics in an adult bariatric-surgery cohort, enabling direct observation of within-person changes over time. We integrated repeated PFAS measurements with detailed assessments of body composition and insulin sensitivity using DXA and IVGTT. The cohort was also predominantly female and majority African American, a population often underrepresented in PFAS research, providing insights into a group with high obesity prevalence^40^. However, important limitations warrant caution. Our sample size was small, which limited our ability to generalize to men and resulted in fewer postoperative observations due to the COVID-19 pandemic. Given the exploratory nature of this work, Spearman correlations were left unadjusted. Furthermore, while it would have been ideal to utilize adjusted mixed-effects models or multiple testing to more robustly evaluate clinical changes over time, the restricted cohort size and the confounding nature of certain categorical variables precluded this approach. Correlational analyses cannot rule out residual confounding by diet, renal clearance, medication use, or other metabolic shifts accompanying weight loss. Also, we analyzed only 3 major PFAS species in which annotation was validated by tandem MS in our workflow. It is possible that other PFAS species would show other relationships to pre- and postoperative body composition and glycemic indexes.

## Conclusions

In summary, this pilot study provides preliminary data that bariatric surgery in adults may coincide with compound-specific changes in circulating PFAS and with baseline differences in body composition. PFHxS declined significantly after surgery, while PFNA and PFOS did not, and higher concentrations of PFNA and PFOS at baseline were inversely related to lean mass, with PFNA also inversely related to body weight. Average PFNA was positively correlated with postoperative changes in HOMA-IR and total lean mass. In supplementary ANCOVA, PFNA was not significantly associated with postoperative outcomes. Although the small sample size and exploratory design limit inference, the longitudinal nature of this work offers a window into PFAS dynamics during rapid weight loss. These findings underscore the value of integrating environmental exposures into bariatric research and highlight the need for larger, time-resolved studies to clarify the role of PFAS in shaping surgical recovery and metabolic outcomes.

## Abbreviations

AIRg: acute insulin response to glucose;
ANCOVA: analysis of covariance;
BMI: body mass index;
CI: confidence interval;
CRC: Clinical Research Center;
DI: disposition index;
DXA: dual-energy X-ray absorptiometry;
HOMA-IR: Homeostatic Model Assessment of Insulin Resistance;
IQR: interquartile range;
IVGTT: intravenous glucose tolerance test;
LC-MS: liquid chromatography–mass spectrometry;
LOD: limit of detection;
MS/MS: tandem mass spectrometry;
PFAS: per- and polyfluoroalkyl substances;
PFHxS: perfluorohexanesulfonic acid;
PFNA: perfluorononanoic acid;
PFOS: perfluorooctanesulfonic acid;
RYGB: Roux-en-Y gastric bypass;
SD: standard deviation;
SE: standard error;
SI: sensitivity index;
UAB: University of Alabama at Birmingham

## Acknowledgements

The authors acknowledge the assistance of the CRC staff in the conduct of this study. Artificial Intelligence tools were used to support English-language editing and debugging statistical code. All AI-generated output was reviewed, verified, and approved by the authors, who take full responsibility for the integrity of the work.

## Author Contributions

**Saketh Sankara:** Conceptualization, Data Curation, Formal Analysis, Investigation, Methodology, Writing – original draft, Visualization, Software

**Matthew R. Smith:** Conceptualization, Data Curation, Formal Analysis, Investigation, Methodology, Software, Supervision, Validation, Writing – review & editing

**Stephanie M. Eick:** Data Curation, Formal Analysis, Software, Methodology, Supervision, Writing – review & editing

**Damaskini Valvi:** Conceptualization, Writing – review & editing

**Tasha M. Burley:** Investigation, Writing – review & editing

**Douglas I. Walker:** Methodology, Writing – review & editing

**Edward Lin:** Investigation, Writing – review & editing

**Elizabeth M. Hechenbleikner:** Investigation, Writing – review & editing

**Lucia A. Gonzalez Ramirez:** Data Curation, Software, Supervision, Writing – review & editing

**Paula-Dene C. Nesbeth:** Data Curation, Writing – review & editing

**Priyathama Vellanki:** Investigation, Methodology, Writing – review & editing

**Barbara A. Gower:** Formal Analysis, Investigation, Writing – review & editing

**Rob McConnell:** Conceptualization, Funding Acquisition, Writing – review & editing

**Dean P. Jones:** Conceptualization, Data Curation, Methodology, Resources, Writing – review & editing

**Jessica A. Alvarez:** Conceptualization, Investigation, Methodology, Writing – review & editing

**Vaia Lida Chatzi:** Conceptualization, Formal Analysis, Funding Acquisition, Methodology, Resources, Writing – review & editing

**Thomas R. Ziegler:** Conceptualization, Data Curation, Formal Analysis, Funding Acquisition, Investigation, Methodology, Visualization, Project Administration, Resources, Supervision, Writing – original draft, Writing – review & editing

## Data Availability Statement

The datasets generated and analyzed during the current study are available from the corresponding author upon reasonable request.

## Supplementary Material

**Table S1.** Longitudinal Plasma Concentrations of PFAS (ng/mL)

| PFAS Species | n | Time Point | Geometric Mean [95% CI] | SD | 50<sup>th</sup> % | 75<sup>th</sup> % | n (%) Below LOD |
| --- | --- | --- | --- | --- | --- | --- | --- |
| PFHxS | 32 | Baseline | 2.353<br>[1.660, 3.334] | 2.630 | 1.952 | 4.391 | 0 (0%) |
|  | 22 | Follow Up | 0.904<br>[0.466, 1.753] | 4.455 | 0.855 | 2.813 | 1 (4.54%) |
|  | 22 | Average | 1.449<br>[0.853, 2.464] | 3.309 | 1.327 | 3.187 | 0 (0%) |
| PFNA | 32 | Baseline | 1.207<br>[0.907, 1.606] | 2.210 | 1.164 | 1.551 | 0 (0%) |
|  | 22 | Follow Up | 0.898<br>[0.604, 1.335] | 2.445 | 0.969 | 1.192 | 1 (4.54%) |
|  | 22 | Average | 1.024<br>[0.750, 1.398] | 2.019 | 0.952 | 1.339 | 0 (0%) |
| PFOS | 32 | Baseline | 7.884<br>[5.814, 10.69] | 2.328 | 6.980 | 12.87 | 0 (0%) |
|  | 22 | Follow Up | 5.619<br>[3.607, 8.753] | 2.718 | 5.250 | 8.150 | 0 (0%) |
|  | 22 | Average | 6.169<br>[4.130, 9.217] | 2.473 | 5.129 | 8.731 | 0 (0%) |

Wilcoxon signed-rank tests for paired pre- and postoperative samples. LOD = limit of detection. n = 22 for all compounds.

**Figure S1.**
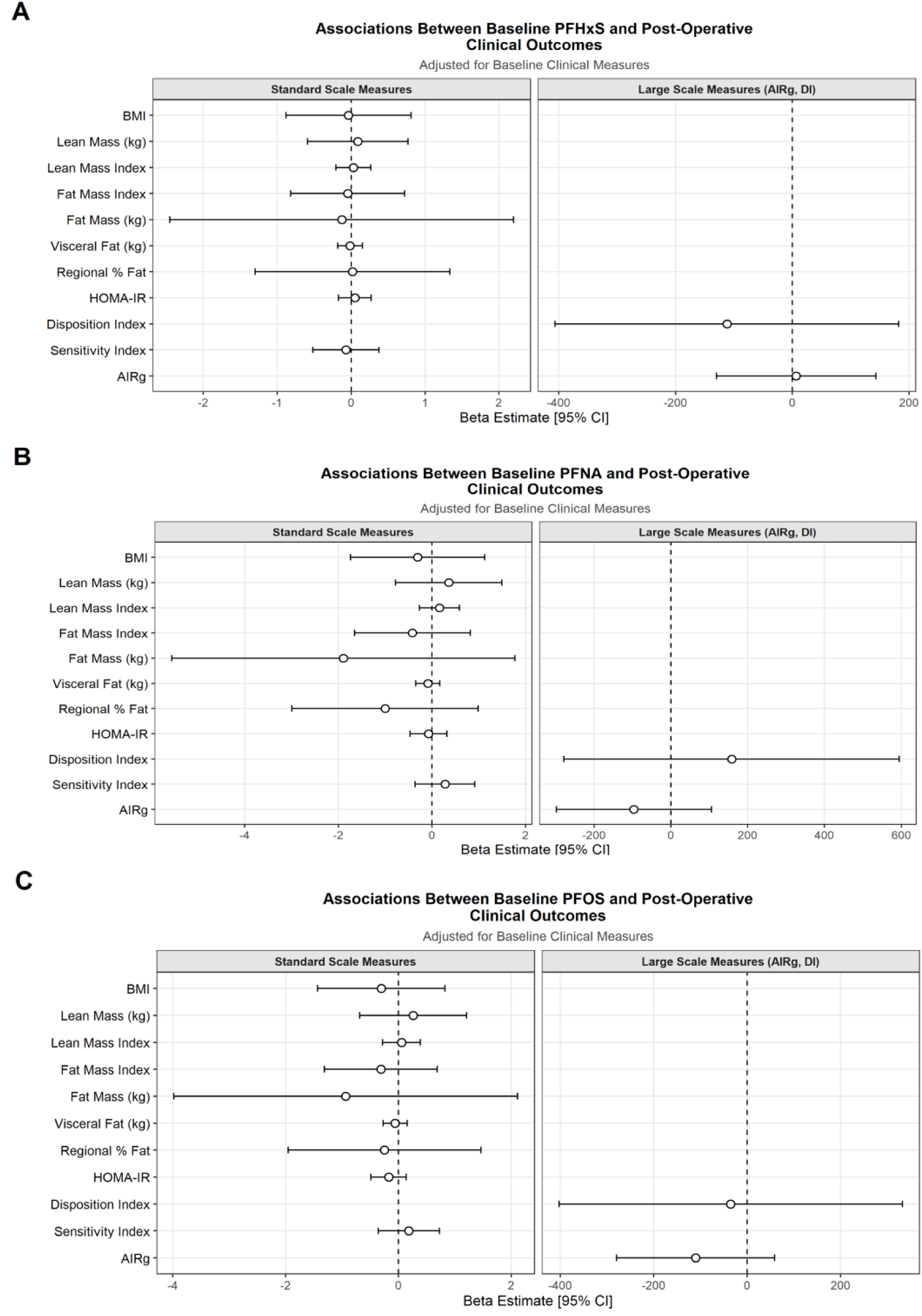
Baseline PFAS ANCOVA with Postoperative Clinical Outcomes. Forest plots display ANCOVA-derived beta estimates from models including baseline PFAS and the corresponding baseline clinical value for (A) PFHxS, (B) PFNA, and (C) PFOS (n=26 for all outcomes except n=23 for AIRg, DI, and SI), respectively. No statistically significant associations or consistent patterns were detected across the analyzed clinical outcomes. AIRg = Acute Insulin Response to Glucose. DI = Disposition Index. HOMA-IR = Homeostatic Model Assessment of Insulin Resistance. SI = Sensitivity Index.

